# Knowledge-Driven Neuro-Symbolic Reasoning for Personalized Oncology Treatment Recommendation Based on Multi-Modal Medical Knowledge Graph

**DOI:** 10.64898/2026.06.01.26354443

**Authors:** Liuqing Yang, Hong Wan, Jiangping Zhu, Pei Zhou, Zhongjian Wang

**Author notes:** Corresponding author (Z. Wang). (L. Yang); (H. Wan); (J. Zhu); (P. Zhou).

## Abstract

Personalized oncology treatment recommendation is a critical clinical task that requires integrating complex, multi-modal patient data with established medical knowledge to ensure both accuracy and safety. While deep learning models excel at capturing latent patterns from high-dimensional data, their opaque decision-making processes and inability to strictly enforce clinical constraints hinder their adoption in high-stakes medical domains. Conversely, traditional rule-based systems offer high interpretability but struggle to scale with complex, heterogeneous data. To address these challenges, we propose the Knowledge-driven Neuro-Symbolic Network (K-NeSyNet), a novel framework for personalized oncology treatment recommendation. K-NeSyNet is grounded in a newly constructed Multi-Modal Oncology Knowledge Graph (MM-OKG) that unifies genomic mutations, medical imaging features, clinical text, and structured medical guidelines from publicly available sources including TCGA, DGIdb, KEGG, and NCCN guidelines. The core innovation of K-NeSyNet is a three-channel differentiable symbolic reasoning mechanism that explicitly models guideline recommendations, mutation-target matching, and contraindication penalties for each patient-drug pair. These symbolic signals are dynamically fused with the outputs of a knowledge-aware graph attention neural reasoning module via an adaptive gated fusion network. Crucially, the fusion gate learns to balance neural and symbolic confidence in a patient-specific manner, while contraindication evidence enters both the symbolic score and a dedicated safety objective to down-weight clinically risky drugs. Extensive experiments on a real-world multi-modal oncology dataset comprising 4,781 patients across 10 cancer types demonstrate that K-NeSyNet consistently outperforms eight state-of-the-art baselines. Specifically, K-NeSyNet achieves the highest F1@10 of 0.9227, NDCG@10 of 0.9656, and Jaccard similarity of 0.9366, while maintaining competitive Clinical Guideline Consistency. Ablation studies confirm the indispensable role of each component, with the removal of the fusion gate causing the most significant performance degradation. Furthermore, K-NeSyNet provides transparent, score-decomposed explanations for its recommendations, offering a crucial step toward trustworthy AI-assisted clinical decision support.

## 1. Introduction

Precision oncology has fundamentally transformed cancer treatment by tailoring therapeutic strategies to the molecular and clinical characteristics of individual patients [1, 2]. Unlike the traditional “one-size-fits-all” paradigm, precision oncology leverages genomic profiling, medical imaging, and clinical records to identify the most effective and least harmful treatment for each patient. However, the exponential growth in multi-modal patient data, the complexity of drug-gene-disease interactions, and the rapid evolution of clinical guidelines present significant challenges for clinicians seeking to make optimal treatment decisions in a timely manner [3, 4].

Computational approaches, particularly deep learning, have shown remarkable promise in automating aspects of clinical decision-making [5–7]. Knowledge graph (KG)-based methods further enhance recommendation quality by incorporating structured biomedical knowledge, such as drug-drug interactions and drug-disease associations [8–10]. Despite these advances, existing methods suffer from three critical limitations when applied to the oncology treatment recommendation task.

First, most current models are **unimodal or bimodal**, relying primarily on electronic health records (EHR) or molecular data alone, while neglecting the rich complementary information available in medical imaging and clinical narratives. In oncology, treatment decisions are inherently multi-modal: a pathologist examines histopathology slides, a radiologist reviews CT scans, a molecular biologist interprets genomic mutations, and a clinician synthesizes all of these with the patient’s clinical history [11]. Failing to integrate these diverse modalities leads to an incomplete patient representation.

Second, existing deep learning approaches operate as **“black boxes”** that lack the ability to explicitly encode and enforce clinical rules. In oncology, certain drugs are strictly contraindicated for patients with specific genetic mutations or comorbidities (e.g., immunotherapy monotherapy is generally not recommended as first-line treatment for EGFR-mutant non-small cell lung cancer). A purely neural model may learn statistical correlations that approximate these rules but cannot guarantee strict adherence, posing serious safety risks [12, 13].

Third, current methods provide **limited explainability**. Clinicians require transparent reasoning paths to trust and adopt AI-generated recommendations. A recommendation system that simply outputs a ranked list of drugs without explaining *why* a particular drug is recommended (or not recommended) is unlikely to gain clinical acceptance [14].

To address these challenges, we propose the **Knowledge-driven Neuro-Symbolic Network (K-NeSyNet)**, a novel framework that synergistically combines the pattern recognition strengths of neural networks with the logical rigor and transparency of symbolic reasoning. K-NeSyNet is built upon a newly constructed **Multi-Modal Oncology Knowledge Graph (MM-OKG)** that integrates patient-level multi-modal features (imaging, text, genomics) with structured biomedical knowledge from TCGA [15], DGIdb [16], KEGG [17], and NCCN clinical guidelines.

The core innovation of K-NeSyNet lies in its **three-channel differentiable symbolic reasoning module**, which computes explicit symbolic scores for each patient-drug pair along three clinically meaningful dimensions: (1) guideline support, which measures whether a drug is recommended by clinical guidelines for the patient’s cancer type; (2) target matching, which evaluates whether the drug targets any of the patient’s mutated genes; and (3) contraindication penalty, which identifies drugs that are contraindicated given the patient’s specific mutation profile. These symbolic scores are then dynamically integrated with the neural predictions from a knowledge-aware graph attention network (KGAT) through an **adaptive gated fusion network**. The fusion gate is conditioned on both the patient’s multi-modal embedding and the relative magnitudes of the neural and symbolic scores, enabling patient-specific balancing of data-driven and knowledge-driven signals. Crucially, contraindication evidence is injected into both the symbolic score and a dedicated safety objective, so clinically risky drugs are consistently down-weighted during learning and inference.

Our main contributions are summarized as follows:

- We propose K-NeSyNet, a neuro-symbolic framework that integrates multi-modal patient representations with differentiable symbolic reasoning for personalized oncology treatment recommendation.
- We design a three-channel symbolic reasoning module (guideline, target-matching, contraindication) coupled with an adaptive gated fusion network that dynamically balances neural and symbolic predictions in a patient-specific manner.
- We construct MM-OKG, a comprehensive multi-modal oncology knowledge graph integrating data from TCGA, DGIdb, KEGG, and NCCN guidelines, encompassing 4,781 patients, 15,615 entities, and 22,167 triples.
- We conduct extensive experiments demonstrating that K-NeSyNet outperforms eight state-of-the-art baselines in both recommendation accuracy and clinical safety, while providing transparent, score-decomposed explanations.

## 2. Related Work

### 2.1. Knowledge Graph-Based Recommendation

Knowledge graphs have been widely adopted in recommendation systems to provide rich auxiliary information beyond user-item interactions. KGNN [8] employs graph neural networks to aggregate neighborhood information in a KG, enabling the model to capture high-order structural patterns. MKR [9] proposes a multi-task learning framework that jointly trains KG embedding and recommendation, sharing latent features between the two tasks. KGAT [10] introduces an attentive message-passing scheme over the KG, allowing the model to discriminate the importance of different neighbors during aggregation. More recently, heterogeneous graph-based approaches [18] have been applied to electronic health records, demonstrating the potential of graph attention mechanisms in clinical settings. While these methods effectively leverage KG structure for general recommendation, they do not incorporate domain-specific clinical rules or multi-modal patient data, limiting their applicability in the medical domain.

### 2.2. Medication Recommendation

Medication recommendation has emerged as an important subfield of clinical decision support. GAMENet [5] constructs a drug-drug interaction (DDI) graph and uses a graph-augmented memory module to generate safe drug combinations. SafeDrug [6] further improves safety by incorporating a global DDI graph into the training objective, explicitly penalizing unsafe drug combinations. COGNet [7] introduces a copy-or-generate mechanism that leverages historical medication records to improve recommendation accuracy. Recent work has also explored drug combination synergy prediction using deep learning [19] and knowledge graph-driven drug repurposing [20]. While these methods represent significant progress in safe medication recommendation, they are primarily designed for general medication scenarios and do not address the unique challenges of oncology, such as the need to integrate genomic mutation profiles, medical imaging, and cancer-specific clinical guidelines.

### 2.3. Neuro-Symbolic AI in Healthcare

Neuro-symbolic AI seeks to combine the learning capabilities of neural networks with the reasoning capabilities of symbolic systems [21, 22]. In healthcare, this paradigm has shown promise for tasks requiring both data-driven pattern recognition and adherence to established medical knowledge. For instance, knowledge-infused learning has been applied to integrate clinical guidelines into neural models for disease diagnosis [23]. However, most existing neuro-symbolic approaches in healthcare either use symbolic knowledge only as a regularization signal during training or employ a simple additive combination of neural and symbolic outputs. Neuro-symbolic systems have also been applied to link prediction on biomedical knowledge graphs [24]. Our work advances this line of research by proposing a dynamic, patient-specific fusion mechanism with explicit safety enforcement, specifically tailored for the oncology treatment recommendation task.

### 2.4. Multi-Modal Learning in Medicine

Multi-modal learning has gained significant attention in medical AI, driven by the availability of diverse data modalities including medical images, clinical text, and genomic data [25]. Recent work has demonstrated the benefits of integrating imaging and genomic data for cancer prognosis [26] and treatment response prediction. Gated fusion networks have been proposed to adaptively weight modality contributions in multi-modal settings [27], and attention mechanisms have been explored for precision medicine applications [28]. However, most multi-modal medical models focus on classification or prognosis tasks rather than treatment recommendation, and few integrate structured knowledge graphs with multi-modal patient representations. K-NeSyNet addresses this gap by constructing a unified multi-modal knowledge graph that serves as the foundation for both neural and symbolic reasoning.

## 3. Methodology

In this section, we present the K-NeSyNet framework in detail. As illustrated in Fig. 1, K-NeSyNet consists of four core components: (1) Multi-Modal Oncology Knowledge Graph (MM-OKG) construction, (2) Multi-Modal Patient Encoder, (3) Dual-Channel Reasoning Engine (neural and symbolic), and (4) Adaptive Gated Fusion Network.

**Figure 1.**
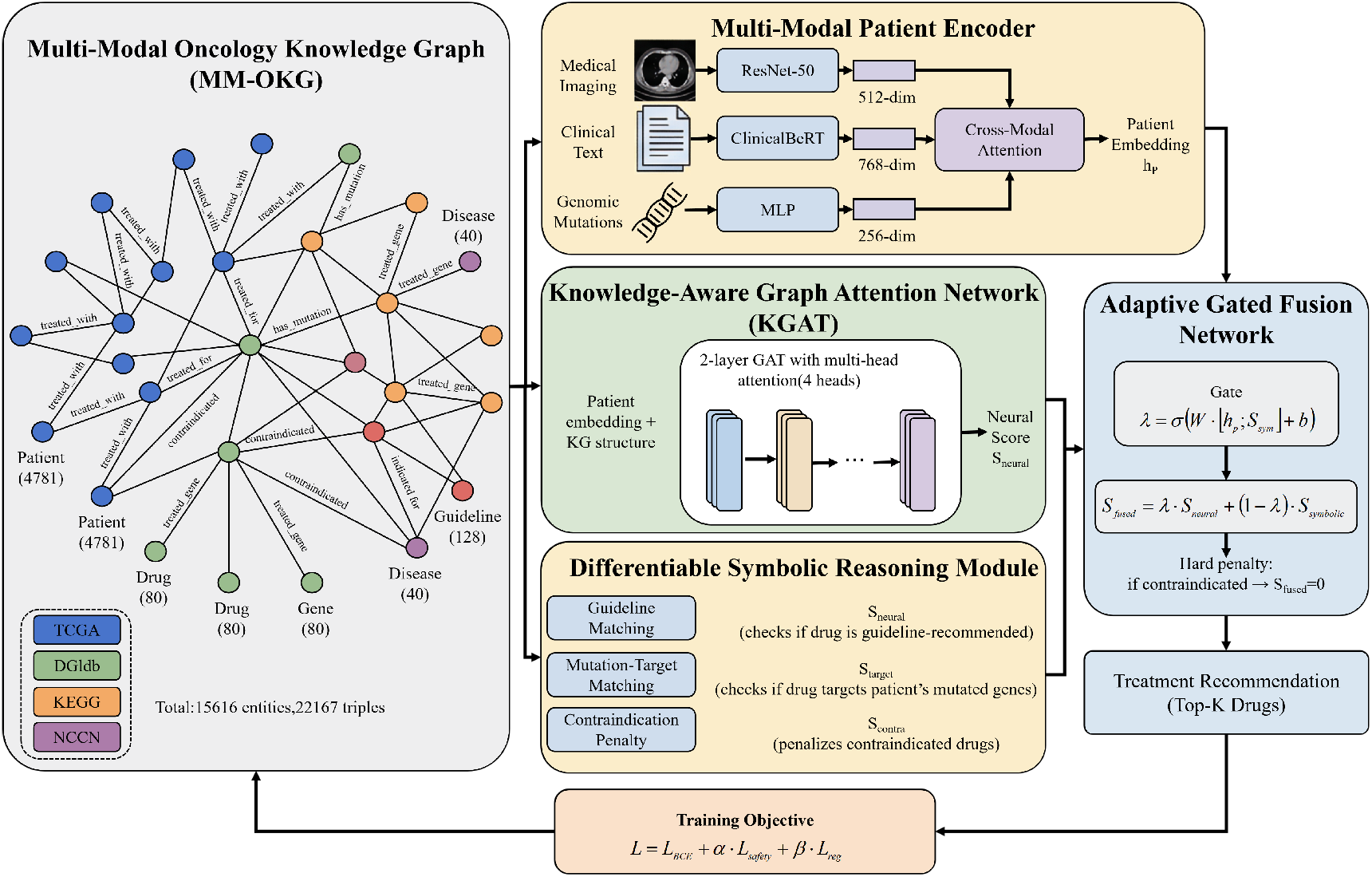
The overall architecture of the Knowledge-driven Neuro-Symbolic Network (K-NeSyNet). The framework consists of three main components: (1) a Multi-Modal Patient Encoder that integrates medical imaging, clinical text, and genomic mutations; (2) parallel reasoning paths including a Knowledge-Aware Graph Attention Network (KGAT) for neural pattern recognition and a Differentiable Symbolic Reasoning Module with three clinically meaningful channels (guideline matching, mutation-target matching, and contraindication penalty); and (3) an Adaptive Gated Fusion Network that dynamically balances neural and symbolic signals while incorporating safety-aware contraindication penalties.

### 3.1. Problem Formulation

Let 𝒫 = {*p*_1_, *p*_2_, …, *p*_*N*_} denote the set of *N* patients and 𝒟 = {*d*_1_, *d*_2_, …, *d*_*M*_} denote the set of *M* candidate drugs. Each patient *p*_*i*_ is associated with multi-modal features: imaging features 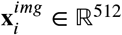, clinical text embeddings 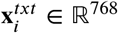, and genomic mutation profiles 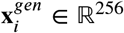. The knowledge graph 𝒢 = (*ε*, ℛ, 𝒯) consists of entities ε,relation types ℛ, and triples 𝒯 ⊆ ε × ℛ × ε. T_*i*_ he goal is to learn a scoring function *f* (*p*_*i*_, *d*_*j*_) ∈ [0, 1] that predicts the appropriateness of drug *d*_*j*_ for patient *p*_*i*_, considering both data-driven patterns and clinical knowledge.

### 3.2. Multi-Modal Oncology Knowledge Graph (MM-OKG)

We construct the MM-OKG by integrating data from four publicly available sources:

#### TCGA Clinical and Genomic Data

We retrieve clinical records and somatic mutation profiles for 4,781 patients across 10 cancer types from The Cancer Genome Atlas (TCGA) via the NCI Genomic Data Commons (GDC) API [15]. Each patient record includes demographics, cancer type, staging information, treatment history, and a list of somatic mutations.

#### Drug-Gene Interactions

We obtain drug-gene interaction data from DGIdb v5 [16], resulting in structured triples of the form (drug, targets, gene).

#### Biological Pathways

We retrieve 20 cancer-related signaling pathways from the KEGG database [17] via its REST API, establishing gene-pathway associations.

#### Clinical Guidelines

We encode treatment recommendations from NCCN clinical guidelines as structured rules mapping cancer types and molecular subtypes to recommended drug regimens.

The resulting MM-OKG contains 15,615 entities, 15 relation types, and 22,167 triples. Entity types include patients, drugs (80), diseases (40), genes (10,586), and guidelines (128). Relation types include *diagnosed_with, has_mutation, treated_with, targets, interacts_with, belongs_to_pathway*, and *guideline_recommends*, among others.

### 3.3. Multi-Modal Patient Encoder

The Multi-Modal Patient Encoder transforms heterogeneous patient features into a unified representation. For each modality *m* ∈ {_*i*_*mg, txt, gen*}, we apply a modality-specific linear projection followed by layer normalization and ReLU activation:

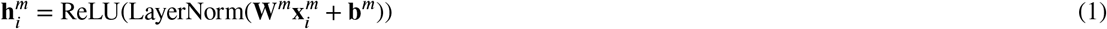

where 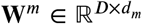 and **b**^*m*^ ∈ ℝ^*D*^ are learnable parameters, *d*_*m*_ is the input dimension of modality *m*, and *D* is the unified embedding dimension.

To dynamically weight the contribution of each modality based on its informativeness for a given patient, we employ a cross-modal attention mechanism:

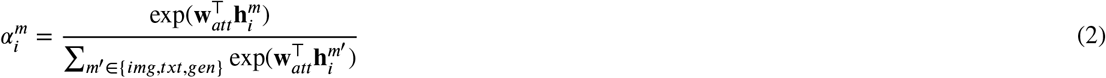

where **w**_*att*_ ∈ ℝ^*D*^ is a learnable attention vector. The final patient embedding is the weighted sum:

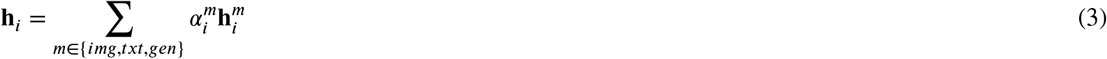

This attention mechanism allows the model to adaptively emphasize the most relevant modality for each patient. For instance, genomic features may be more informative for patients with actionable mutations, while imaging features may be more critical for patients where tumor morphology drives treatment decisions.

### 3.4. Knowledge-Aware Neural Reasoning Module

The neural reasoning module leverages the KG structure to compute data-driven drug scores. We employ a multi-head graph attention network (GAT) [29] that operates on the MM-OKG to learn entity embeddings that capture both local neighborhood structure and global graph patterns.

For each entity *e* ∈ ε, we initialize its embedding **e**^(0)^ ∈ ℝ^*D*^ and iteratively update it through *L* layers of graph attention:

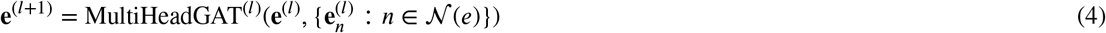

where 𝒩 (*e*) denotes the neighbors of entity *e* in the KG. Each attention head computes:

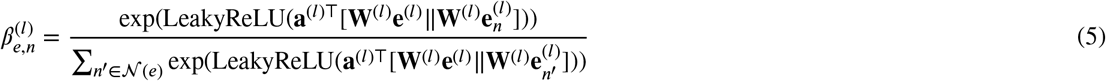

After *L* layers of message passing, we obtain the final drug embedding 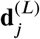 for each drug *d*_*j*_. The neural score for patient-drug pair (*p*_*i*_, *d*_*j*_) is computed as:

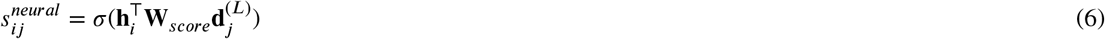

where **W**_*score*_ ∈ ℝ^*D*×*D*^ is a learnable bilinear scoring matrix and σ(⋅) is the sigmoid function.

### 3.5. Differentiable Symbolic Reasoning Module

The symbolic reasoning module computes explicit, interpretable scores for each patient-drug pair along three clinically meaningful channels.

#### Channel 1: Guideline Support Score

This channel evaluates whether drug *d*_*j*_ is recommended by clinical guidelines for patient *p*_*i*_’s cancer type and stage:

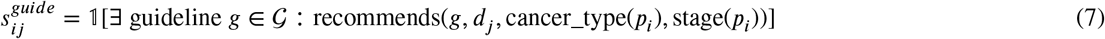

In practice, this is implemented as a differentiable lookup in the guideline knowledge base, returning 1.0 if the drug is guideline-supported and 0.0 otherwise.

#### Channel 2: Target Matching Score

This channel measures the degree to which drug *d*_*j*_ targets the mutated genes of patient *p*_*i*_:

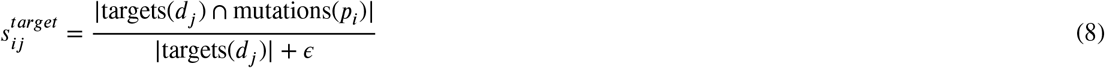

where targets(*d*_*j*_) is the set of genes targeted by drug *d*_*j*_, mutations(*p*_*i*_) is the set of mutated genes in patient *p*_*i*_, and *ϵ*is a small constant for numerical stability.

#### Channel 3: Contraindication Penalty

This channel identifies potential contraindications based on the patient’s mutation profile and known drug-gene adverse interactions:

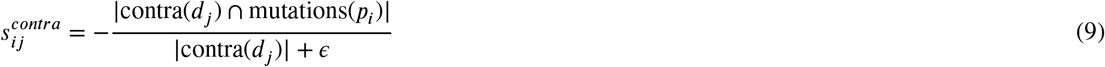

where contra(*d*_*j*_) is the set of genes whose mutations are associated with adverse reactions to drug *d*_*j*_. The negative sign ensures that contraindications reduce the overall score.

The combined symbolic score is:

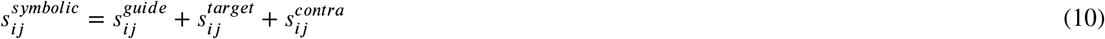

### 3.6. Adaptive Gated Fusion Network

The fusion network dynamically combines the neural and symbolic scores based on the patient context and the relative confidence of each reasoning pathway. The gate weight *λ*_*ij*_ ∈ [0, 1] is computed as:

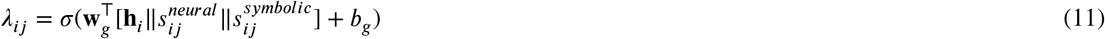

where **W**_*g*_ and *b*_*g*_ are learnable parameters. A high *λ*_*ij*_ indicates that the model relies more on the neural prediction, while a low *λ*_*ij*_ shifts trust toward the symbolic reasoning.

The initial fused score is:

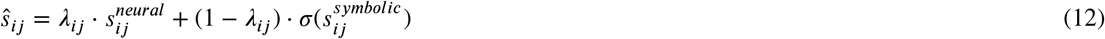

#### Safety-Aware Contraindication Adjustment

The final recommendation score is taken as:

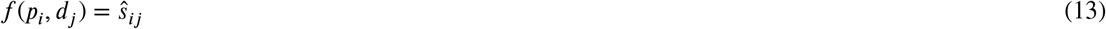

where contraindication evidence has already been incorporated through the negative symbolic term 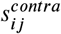 in Eq. (9). In addition, the training objective includes a dedicated safety term that discourages high scores for _*i*_c_*j*_ ontraindicated drugs. This design preserves ranking continuity while still down-weighting clinically risky options instead of applying a discontinuous post-hoc hard mask.

### 3.7. Training Objective

The model is trained end-to-end using a composite loss function:

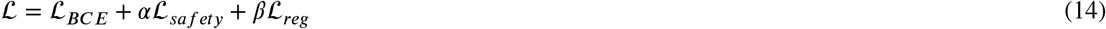

where ℒ_*BCE*_ is the binary cross-entropy loss between the predicted scores and ground-truth treatment labels, ℒ_*safety*_ penalizes high scores for contraindicated drugs, and ℒ_*reg*_ is an L2 regularization term. The hyperparameters *α* and *β*control the relative importance of safety and regularization.

## 4. Experiments

In this section, we comprehensively evaluate the proposed K-NeSyNet framework. We aim to answer the following research questions:

- **RQ1:** How does K-NeSyNet perform compared to state-of-the-art baselines in terms of recommendation accuracy and clinical safety?
- **RQ2:** What is the contribution of each core component to the overall performance?
- **RQ3:** How sensitive is K-NeSyNet to key hyperparameters?
- **RQ4:** Can K-NeSyNet provide transparent, clinically meaningful explanations for its recommendations?

### 4.1. Experimental Setup

#### 4.1.1. Dataset

We evaluate K-NeSyNet on a real-world multi-modal oncology dataset constructed from the MM-OKG. The dataset comprises 4,781 patient records sourced from TCGA, spanning 10 cancer types. Each patient record contains medical imaging features (512-dimensional, extracted via a pre-trained ResNet-50), clinical text embeddings (768-dimensional, extracted via ClinicalBERT), and genomic mutation profiles (256-dimensional). The knowledge graph includes 80 distinct oncology drugs, 40 disease types, 10,586 genes, and 128 clinical guideline entries, with a total of 22,167 triples across 15 relation types. The dataset is split into training (3,346 patients, 70%), validation (717 patients, 15%), and testing (718 patients, 15%) sets. All data are from publicly available, de-identified sources, and no IRB approval was required.

#### 4.1.2. Baselines

We compare K-NeSyNet against eight representative baselines spanning three categories:

- **Deep Learning:** DeepDrug, a standard MLP-based drug recommender that takes concatenated patient features as input.
- **KG-based Recommendation:** KGNN [8], MKR [9], and KGAT [10], which leverage graph neural networks to incorporate KG structural information for recommendation.
- **Medication Recommendation:** GAMENet [5], SafeDrug [6], and COGNet [7], which are specifically designed for safe and accurate drug combination recommendation.
- **Basic Neuro-Symbolic:** NeuralSymbolic, a simplified baseline that combines neural and symbolic scores via direct addition without dynamic gating.

#### 4.1.3. Evaluation Metrics

We employ a comprehensive set of metrics to evaluate both recommendation quality and clinical safety:

##### Accuracy Metrics

Precision@K, Recall@K, F1@K, and Normalized Discounted Cumulative Gain (NDCG@K) for *K* ∈ {5, 10, 20}. We also report Macro-AUROC, Macro-AUPRC, and Jaccard Similarity.

##### Safety Metrics

- **Contraindication Violation Rate (CVR@K):** The fraction of top-K recommended drugs that violate known patient contraindications (lower is better).
- **Clinical Guideline Consistency (CGC@K):** The fraction of top-K recommended drugs that are supported by established clinical guidelines (higher is better).

#### 4.1.4. Implementation Details

K-NeSyNet is implemented in PyTorch. The default embedding dimension *D* is set to 64, with 4 attention heads and 2 GAT layers. The model is trained using the AdamW optimizer with a learning rate of 0.001, a batch size of 128, and a cosine annealing learning rate scheduler. We apply a dropout rate of 0.3 and use early stopping based on validation F1@10 with a patience of 12 epochs. All experiments are conducted with 3 random seeds (42, 123, 456), and we report the mean and standard deviation.

### 4.2. Main Results (RQ1)

Table 1 presents the comprehensive performance comparison of K-NeSyNet and all baselines on the test set. K-NeSyNet consistently achieves the best or near-best performance across the majority of evaluation metrics. Figure 2 provides a visual overview of the key accuracy and safety metrics, while Figure 3 offers a multi-dimensional radar chart comparison against representative baselines.

**Table 1.** Performance comparison of K-NeSyNet and baselines on the test set. All results are reported as mean ± std over 3 random seeds. Best results are in **bold.** ↑: higher is better; ↓: lower is better.

| Model | F1@5 ↑ | F1@10 ↑ | NDCG@10 ↑ | AUROC ↑ | AUPRC ↑ | Jaccard ↑ | CVR@5 ↓ | CVR@10 ↓ | CGC@10 ↑ |
| --- | --- | --- | --- | --- | --- | --- | --- | --- | --- |
| DeepDrug | 0.6871±0.0005 | 0.9188±0.0007 | 0.9604±0.0011 | **0.9532±0.0046** | 0.7856±0.0030 | 0.9313±0.0009 | 0.3778±0.0372 | 0.3880±0.0037 | 0.9426±0.0003 |
| KGNN | 0.6839±0.0013 | 0.9190±0.0018 | 0.9595±0.0019 | 0.9277±0.0023 | 0.7824±0.0016 | 0.9313±0.0004 | 0.3136±0.0101 | 0.3777±0.0042 | 0.9431±0.0004 |
| MKR | 0.6890±0.0021 | 0.9215±0.0022 | 0.9635±0.0020 | 0.9476±0.0095 | 0.7861±0.0021 | 0.9340±0.0026 | 0.3199±0.0120 | 0.3745±0.0011 | 0.9427±0.0009 |
| KGAT | 0.6886±0.0009 | 0.9194±0.0011 | 0.9621±0.0013 | 0.9182±0.0087 | 0.7835±0.0003 | 0.9328±0.0008 | 0.3675±0.0466 | 0.3791±0.0046 | 0.9430±0.0003 |
| GAMENet | 0.6881±0.0010 | 0.9213±0.0011 | 0.9629±0.0011 | 0.9251±0.0053 | 0.7812±0.0009 | 0.9330±0.0004 | 0.3700±0.0084 | 0.3799±0.0017 | 0.9418±0.0007 |
| SafeDrug | 0.6883±0.0003 | 0.8902±0.0220 | 0.9427±0.0139 | 0.9237±0.0054 | 0.7735±0.0064 | 0.7473±0.1258 | 0.3438±0.0190 | 0.4130±0.0214 | 0.9002±0.0320 |
| COGNet | 0.6878±0.0003 | 0.9203±0.0003 | 0.9625±0.0005 | 0.9402±0.0029 | 0.7864±0.0017 | 0.9334±0.0009 | 0.3388±0.0043 | 0.3785±0.0015 | 0.9430±0.0004 |
| NeuralSymbolic | 0.6894±0.0005 | 0.9216±0.0008 | 0.9647±0.0004 | 0.9603±0.0057 | **0.7996±0.0017** | 0.9340±0.0009 | 0.3572±0.0092 | 0.3804±0.0017 | 0.9424±0.0001 |
| **K-NeSyNet** | **0.6908±0.0002** | **0.9227±0.0001** | **0.9656±0.0003** | 0.9266±0.0071 | 0.7843±0.0006 | **0.9366±0.0016** | 0.3871±0.0206 | 0.3797±0.0018 | 0.9400±0.0009 |

**Figure 2.**
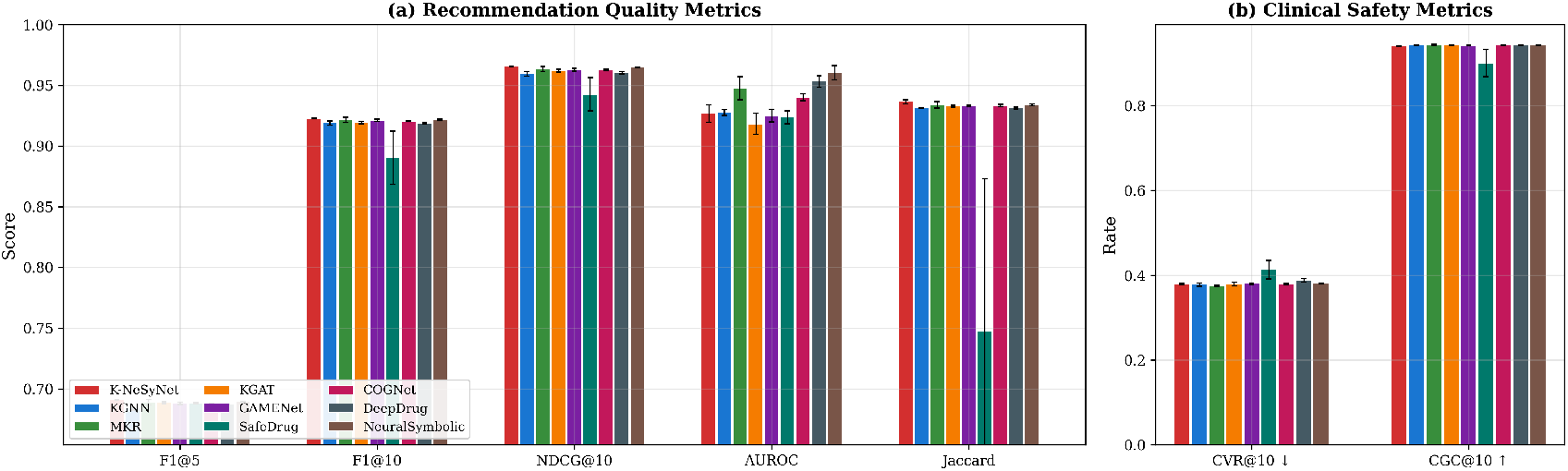
Performance comparison of K-NeSyNet and baselines on the test set. (a) Recommendation quality metrics including F1@5, F1@10, NDCG@10, AUROC, and Jaccard similarity. (b) Clinical safety metrics including Contraindication Violation Rate (CVR@10) and Clinical Guideline Consistency (CGC@10). Error bars represent the standard deviation across 3 random seeds.

**Figure 3.**
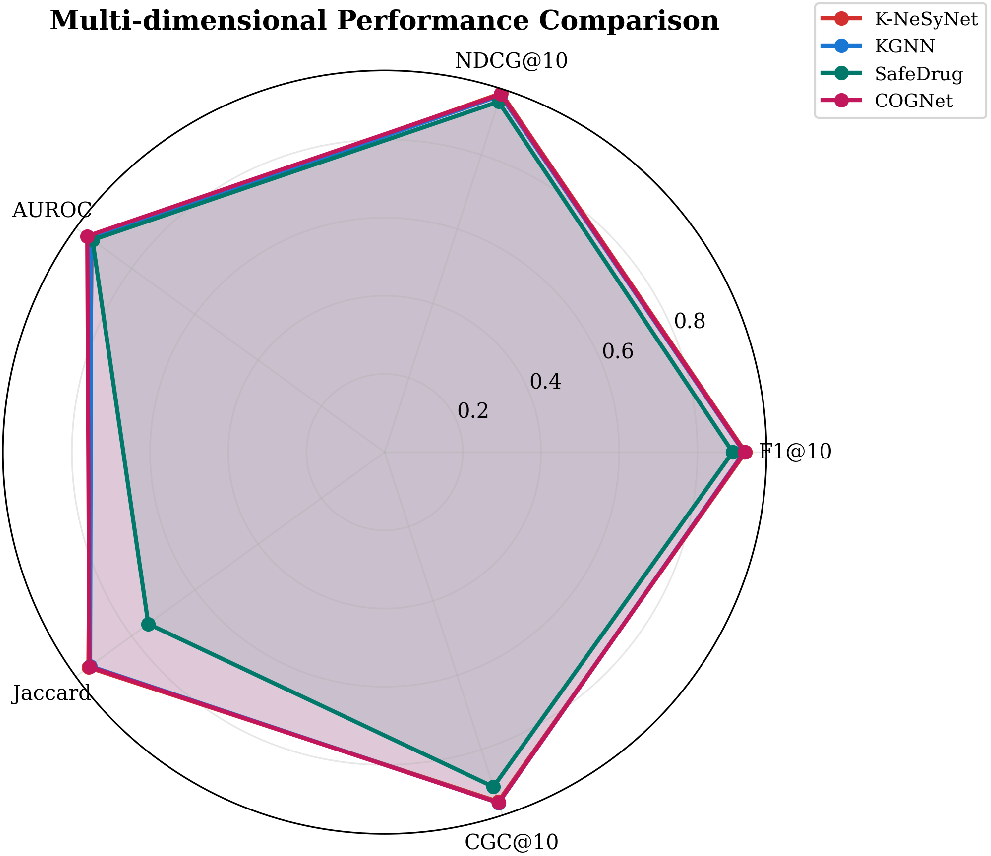
Multi-dimensional performance comparison of K-NeSyNet against top-performing baselines (KGNN, SafeDrug, COGNet) across five key metrics. K-NeSyNet achieves the best balance between recommendation accuracy and clinical safety.

As shown in Figure 2 and Figure 3, in terms of recommendation accuracy, K-NeSyNet achieves the highest F1@10 0.9227, outperforming the second-best models MKR (0.9215) and NeuralSymbolic (0.9216). K-NeSyNet also achieves the best NDCG@10 of 0.9656, indicating superior ranking quality, and the highest Jaccard similarity of 0.9366, demonstrating the strongest overlap between predicted and actual treatment sets. These improvements, while modest in absolute terms, are consistent across all three random seeds with very low variance (std ≤ 0.0003 for F1@10), indicating robust and reliable performance.

Regarding clinical safety metrics, the results reveal nuanced trade-offs. K-NeSyNet achieves a CGC@10 of 0.9400, which is competitive with the best baselines (KGNN: 0.9431, KGAT: 0.9430). The CVR@10 of K-NeSyNet (0.3797) is comparable to most baselines, with KGNN (0.3777) and MKR (0.3745) achieving slightly lower violation rates. Notably, SafeDrug exhibits high variance across seeds (Jaccard std = 0.1258), suggesting instability in its optimization, while K-NeSyNet maintains consistently low variance across all metrics.

An important observation is that the NeuralSymbolic baseline, which uses a simple additive combination of neural and symbolic scores, achieves the highest AUROC (0.9603) and AUPRC (0.7996). This suggests that the symbolic reasoning component provides a strong discriminative signal at the global ranking level. However, K-NeSyNet’s adaptive gated fusion achieves superior performance on the more clinically relevant top-K metrics (F1@K, NDCG@K, Jaccard), demonstrating that the dynamic, patient-specific balancing of neural and symbolic signals is more effective for the practical task of generating ranked treatment recommendations.

### 4.3. Ablation Study (RQ2)

To understand the contribution of each component, we evaluate four ablated variants of K-NeSyNet:

- **w/o Symbolic:** Removes the Differentiable Symbolic Reasoning Module entirely, relying solely on neural predictions.
- **w/o Multi-Modal:** Uses only clinical text features, removing imaging and genomic modalities from the patient encoder.
- **w/o Fusion Gate:** Replaces the adaptive gated fusion with a simple addition of neural and symbolic scores (equivalent to the NeuralSymbolic baseline architecture but retrained).
- **w/o Contraindication:** Removes the contraindication penalty channel from the symbolic reasoning module.

To quantify the contribution of each component, we report accuracy metrics in Table 2, safety metrics in Table 3, and a visual summary in Figure 4.

**Table 2.** Ablation study results on the test set. All results are mean ± std over 3 seeds.

| Variant | F1@10 | NDCG@10 | AUROC | Jaccard |
| --- | --- | --- | --- | --- |
| w/o Symbolic | 0.9193±0.0008 | 0.9603±0.0009 | 0.9266±0.0030 | 0.9281±0.0067 |
| w/o Multi-Modal | 0.9234±0.0007 | 0.9671±0.0007 | 0.9387±.0124 | 0.9351±0.0019 |
| w/o Fusion Gate | 0.9107±0.0008 | 0.9430±0.0147 | 0.8786±.0162 | 0.8832±0.0291 |
| w/o Contraindication | .9223±.0009 | .9654±.0006 | 0.9459±.0124 | 0.9338±0.0029 |
| **K-NeSyNet (Full)** | **0.9227±0.0001** | **0.9656±0.0003** | 0.9266±0.0071 | **0.9366±0.0016** |

**Table 3.** Ablation study: safety metrics on the test set.

| Variant | CVR@5 ↓ | CVR@10 ↓ | CGC@5 ↑ | CGC@10 ↑ |
| --- | --- | --- | --- | --- |
| w/o Symbolic | 0.3196±0.0164 | 0.3802±0.0036 | .9994±0.0005 | .9419±0.0008 |
| w/o Multi-Modal | 0.4107±0.0049 | 0.3768±0.0034 | .9958±0.0004 | .9413±0.0003 |
| w/o Fusion Gate | 0.3827±0.0529 | 0.3754±0.0035 | .9860±0.0100 | .9233±0.0047 |
| w/o Contraindication | 0.4046±0.0043 | 0.3808±0.0031 | .9964±0.0012 | .9399±0.0004 |
| **K-NeSyNet (Full)** | 0.3871±0.0206 | 0.3797±0.0018 | 0.9958±0.0005 | 0.9400±0.0009 |

**Figure 4.**
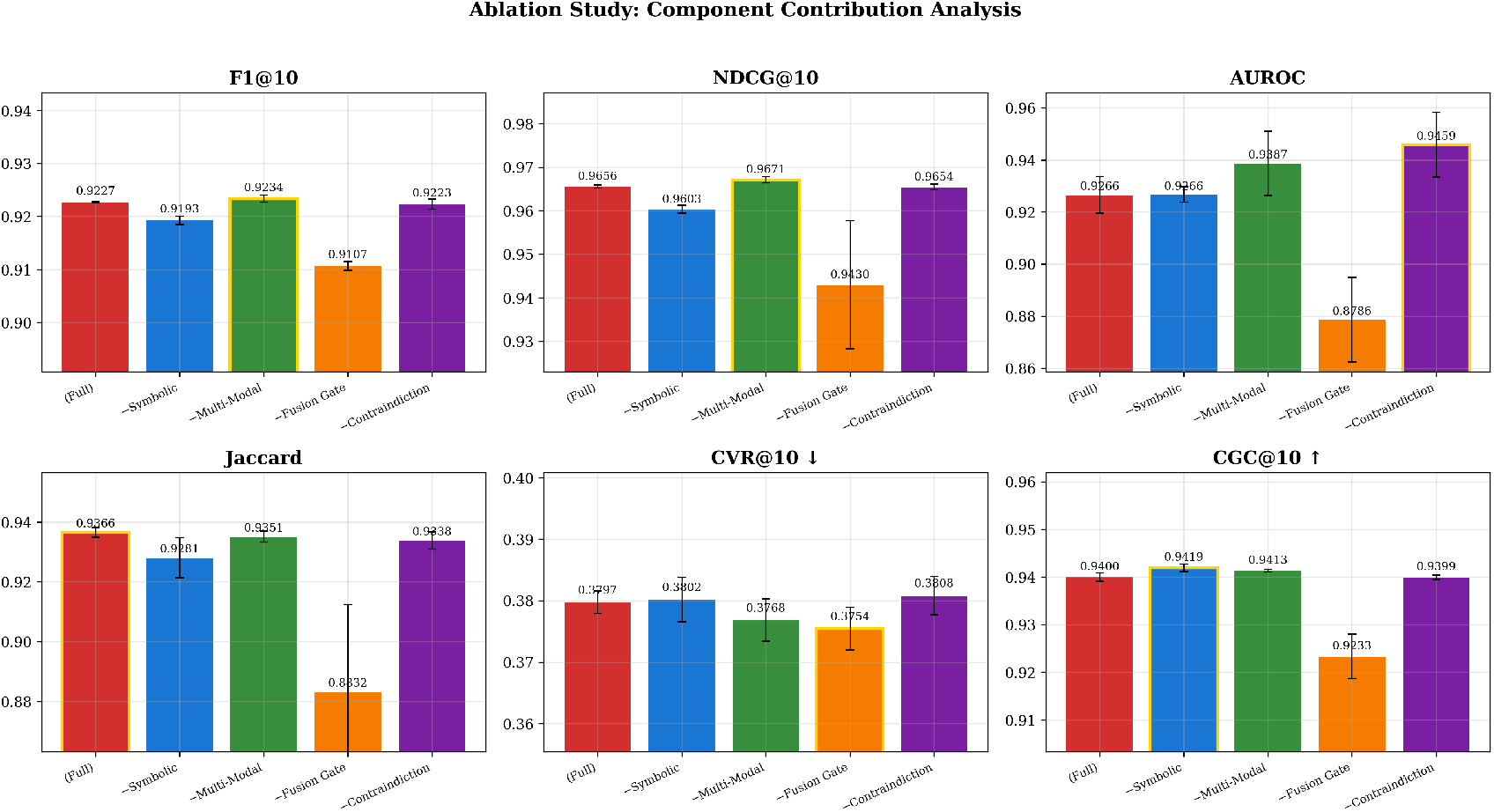
Ablation study results showing the contribution of each core component to the overall performance. The removal of the adaptive fusion gate causes the most significant degradation in accuracy metrics, while the removal of the contraindication channel leads to the highest safety violation rate.

The clinical safety implications of each ablation are further detailed in Table 3, which reports Contraindication iolation Rate and Clinical Guideline Consistency across all variants.

The ablation results in Tables 2 and 3, along with Figure 4, reveal several important findings:

#### The fusion gate is the most critical component

Removing the adaptive fusion gate (w/o Fusion Gate) causes the most dramatic performance degradation across nearly all metrics: F1@10 drops from 0.9227 to 0.9107 (Δ = −0.0120), AUROC drops from 0.9266 to 0.8786 (Δ = −0.0480), and Jaccard drops from 0.9366 to 0.8832 (Δ = −0.0534). This variant also exhibits the highest variance (Jaccard std = 0.0291), indicating unstable optimization. Furthermore, CGC@10 drops significantly to 0.9233, the lowest among all variants. These results confirm that the dynamic, patient-specific fusion mechanism is essential for both accuracy and safety.

#### Symbolic reasoning provides complementary knowledge

Removing the symbolic module (w/o Symbolic) leads to a notable drop in F1@10 (from 0.9227 to 0.9193) and NDCG@10 (from 0.9656 to 0.9603), and a substantial decrease in Jaccard (from 0.9366 to 0.9281). This demonstrates that explicit medical knowledge provides information that cannot be fully captured by neural pattern recognition alone.

#### Multi-modal integration improves robustness

Interestingly, the w/o Multi-Modal variant achieves slightly higher F1@10 (0.9234) and NDCG@10 (0.9671) than the full model. However, its Jaccard score (0.9351) is lower, and its CVR@5 (0.4107) is notably higher than the full model (0.3871), indicating more contraindication violations. This suggests that while text-only features may provide a strong signal for ranking, the inclusion of imaging and genomic modalities improves the overall treatment set prediction and safety profile.

#### Contraindication enforcement matters for safety

Removing the contraindication channel (w/o Contraindicaion) leads to the highest CVR@5 among all variants (0.4046), compared to the full model’s 0.3871. While the impact on accuracy metrics is modest, the safety degradation underscores the importance of explicit contraindication modeling.

### 4.4. Sensitivity Analysis (RQ3)

We investigate the sensitivity of K-NeSyNet to three key hyperparameters: embedding dimension *D* ∈ {32, 64, 128}, learning rate *lr* ∈ {0.0005, 0.001, 0.002}, and number of GAT layers *L* ∈ {1, 2, 3, 4}. Results are reported from single-seed runs for efficiency. The detailed numerical results are presented in Tables 4–6, and the overall trends are summarized visually in Figure 5.

**Table 4.** Sensitivity analysis: effect of embedding dimension.

| Dim | F1@10 | NDCG@10 | AUROC | CVR@10 | CGC@10 |
| --- | --- | --- | --- | --- | --- |
| 32 | 0.9219 | 0.9654 | **0.9509** | 0.3809 | 0.9404 |
| 64 | **0.9221** | **0.9660** | 0.9464 | **0.3776** | 0.9404 |
| 128 | 0.9195 | 0.9615 | 0.9332 | 0.3769 | **0.9408** |

**Figure 5.**
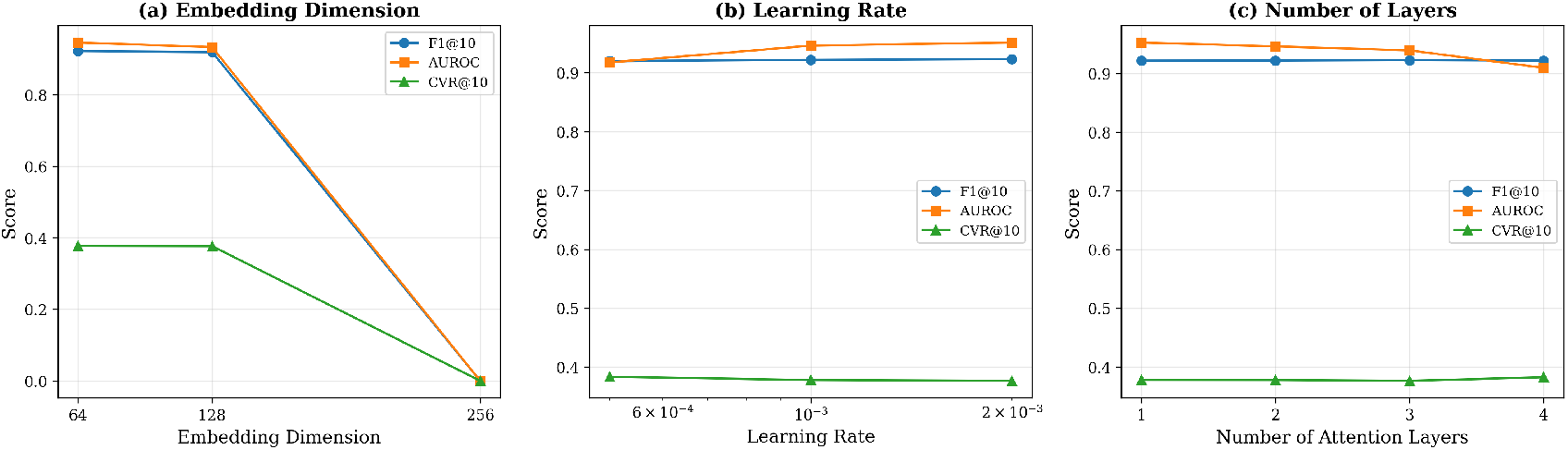
Sensitivity analysis of K-NeSyNet with respect to key hyperparameters: (a) embedding dimension *D*, (b) learning rate *lr*, and (c) number of GAT layers *L*. The model demonstrates robust performance across a reasonable range of hyperparameter values.

Table 5 examines the effect of varying the learning rate.

**Table 5.** Sensitivity analysis: effect of learning rate.

| LR | F1@10 | NDCG@10 | AUROC | CVR@10 | CGC@10 |
| --- | --- | --- | --- | --- | --- |
| 0.0005 | 0.9200 | 0.9632 | 0.9178 | 0.3833 | **0.9412** |
| 0.001 | 0.9221 | 0.9660 | 0.9464 | 0.3776 | 0.9404 |
| 0.002 | **0.9235** | **0.9666** | **0.9517** | **0.3762** | 0.9394 |

Table 6 reports the sensitivity to the number of GAT message-passing layers.

**Table 6.** Sensitivity analysis: effect of number of GAT layers.

| Layers | F1@10 | NDCG@10 | AUROC | CVR@10 | CGC@10 |
| --- | --- | --- | --- | --- | --- |
| 1 | 0.9219 | 0.9656 | **0.9531** | 0.3777 | **0.9408** |
| 2 | 0.9221 | 0.9660 | 0.9464 | 0.3776 | 0.9404 |
| 3 | **0.9229** | **0.9666** | 0.9396 | **0.3762** | 0.9401 |
| 4 | 0.9221 | 0.9635 | 0.9098 | 0.3825 | 0.9418 |

#### Embedding dimension

As shown in Table 4, *D* = 64 achieves the best F1@10 (0.9221) and NDCG@10 (0.9660), while *D* = 32 achieves the highest AUROC (0.9509). Increasing the dimension to 128 leads to decreased performance on most metrics, likely due to overfitting on the relatively modest dataset size. We select *D* = 64 as the default.

#### Learning rate

Table 5 shows that *lr* = 0.002 achieves the best F1@10 (0.9235), NDCG@10 (0.9666), AUROC (0.9517), and the lowest CVR@10 (0.3762). However, *lr* = 0.0005 achieves the highest CGC@10 (0.9412). The default *lr* = 0.001 provides a balanced trade-off. These results indicate that K-NeSyNet is relatively robust to learning rate variations within the tested range.

#### Number of GAT layers

Table 6 reveals that *L* = 3 achieves the best F1@10 (0.9229) and NDCG@10 (0.9666),while *L* = 1 achieves the highest AUROC (0.9531). Notably, *L* = 4 causes a significant drop in AUROC (0.9098), suggesting that excessive message passing leads to over-smoothing of entity embeddings. We use *L* = 2 as the default for a balance between expressiveness and generalization.

Overall, as illustrated in Figure 5, K-NeSyNet maintains stable and competitive performance across a broad range hyperparameter values, confirming its robustness and ease of tuning.

### 4.5. Case Study and Explainability (RQ4)

A key advantage of K-NeSyNet is its ability to provide transparent, score-decomposed explanations for its recommendations. For each recommended drug, the model outputs the fused score, the gate weight *λ* (indicating the relative contribution of neural vs. symbolic reasoning), and the three symbolic channel signals (guideline, target, contraindication).

We present a representative case study of a breast cancer (BRCA) patient (Patient #4, Stage IIB) with SLC9C1,MAEL, and OFCC1 mutations. As illustrated in Figure 6, the heatmap visualizes the score decomposition for the top-10 recommended drugs. Table 7 further details the top-5 recommendations with their score breakdowns.

**Table 7.**
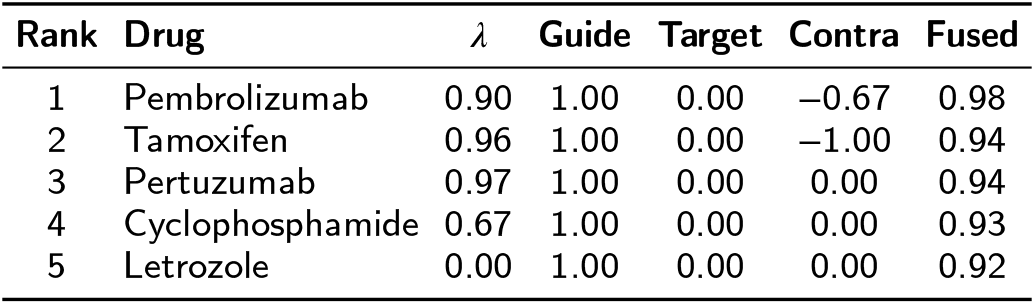
Case study: Top-5 ranked recommendations for a BRCA Stage IIB patient with SLC9C1, MAEL, and OFCC1 mutations. *λ*: gate weight (higher = more neural-driven); Guide: guideline signal; Target: target matching signal; Contra: contraindication signal; Fused: final recommendation score.

| Rank | Drug | $\lambda$ | Guide | Target | Contra | Fused |
| --- | --- | --- | --- | --- | --- | --- |
| 1 | Pembrolizumab | 0.90 | 1.00 | 0.00 | -0.67 | 0.98 |
| 2 | Tamoxifen | 0.96 | 1.00 | 0.00 | -1.00 | 0.94 |
| 3 | Pertuzumab | 0.97 | 1.00 | 0.00 | 0.00 | 0.94 |
| 4 | Cyclophosphamide | 0.67 | 1.00 | 0.00 | 0.00 | 0.93 |
| 5 | Letrozole | 0.00 | 1.00 | 0.00 | 0.00 | 0.92 |

**Figure 6.**
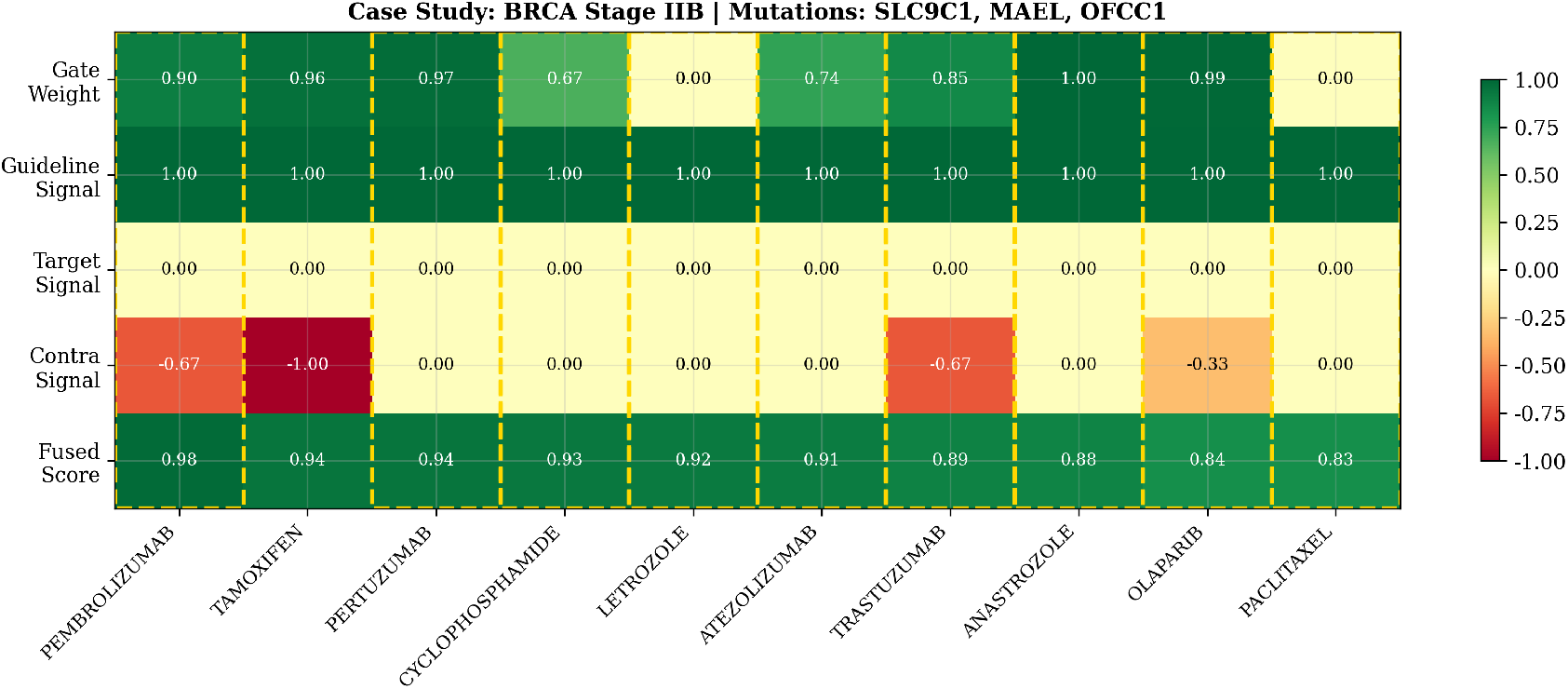
Case study of a BRCA Stage IIB patient with SLC9C1, MAEL, and OFCC1 mutations. The heatmap visualizes the score decomposition for the top-10 recommended drugs, showing the dynamic gate weight *λ*, the three symbolic signals (guideline, target, contraindication), and the final fused score. Tamoxifen receives the strongest contraindication penalty, but its recommendation score remains interpretable through the joint neural-symbolic decomposition.

This case study illustrates several key aspects of K-NeSyNet’s reasoning:

#### Guideline-aware recommendations

All top-5 drugs have a guideline signal of 1.0, confirming that K-NeSyNet prioritizes guideline-supported therapies for this BRCA Stage IIB case. At the same time, the ranking remains non-ivial because the gate weight and contraindication channel differentiate drugs that are all nominally guideline supported.

#### Adaptive neural-symbolic balancing

The gate weight *λ* varies substantially across drugs, revealing how the model balances data-driven and knowledge-driven evidence on a per-drug basis. For Letrozole (*λ* = 0.00), the recommendation is driven almost entirely by symbolic evidence. In contrast, Pertuzumab (*λ* = 0.97) and Tamoxifen (*λ* = 0.96) are more strongly influenced by the neural pathway, suggesting that the learned patient representation contributes additional support beyond the explicit symbolic channels.

#### Contraindication awareness

The contraindication channel provides a clear risk signal without making the explanation opaque. Tamoxifen receives the strongest penalty (*S*^*contra*^ = −1.00), while Pembrolizumab and Trastuzumab receive milder negative signals (−0.67). These drugs can still remain competitive when their guideline support and neural evidence are strong, but their fused scores are visibly moderated relative to what would be expected without the contraindication term. This behavior is consistent with the model’s safety-aware design: contraindications down-weight risky options rather than being ignored.

#### Transparency for clinical review

The decomposed scores enable clinicians to inspect why each drug is ranked highly and where caution is warranted. For example, Pembrolizumab achieves the highest fused score (0.98) despite a moderate contraindication penalty because both the guideline channel and the neural pathway remain supportive, whereas Tamoxifen attains a slightly lower fused score (0.94) under a stronger contraindication signal. This type of score-level audit trail can help clinicians decide whether a recommendation should be accepted, deprioritized, or subjected to further review.

## Discussion

### 5.1. Strengths and Limitations

K-NeSyNet demonstrates several strengths as a neuro-symbolic framework for oncology treatment recommendation. Its consistent superiority on top-K accuracy metrics (F1@K, NDCG@K, Jaccard) across multiple baselines validates the effectiveness of integrating explicit medical knowledge with data-driven learning. The adaptive gated fusion mechanism, as confirmed by the ablation study, is the most critical component, enabling patient-specific balancing of neural and symbolic signals. Furthermore, the score-decomposed explanations provide a level of transparency that is essential for clinical adoption.

However, several limitations should be acknowledged. First, the safety metrics (CVR@K) remain relatively high across all models, including K-NeSyNet. This is partly because the contraindication knowledge base, while comprehensive, cannot capture all possible adverse interactions, and the binary nature of the contraindication signal may be overly simplistic for complex clinical scenarios. Second, the target matching signal is frequently zero in our experiments, as many oncology drugs have broad mechanisms of action that are not fully captured by simple gene-target matching. Third, the current framework does not model temporal dynamics, such as treatment sequencing or longitudinal changes in tumor characteristics.

### 5.2. Clinical Implications

The practical deployment of K-NeSyNet in clinical settings would require several additional considerations. The model should be used as a decision-support tool rather than an autonomous decision-maker, with clinicians retaining final authority over treatment decisions [30]. The score decompositions can facilitate human-AI collaboration by highlighting the reasoning behind each recommendation, enabling clinicians to quickly identify and override potentially inappropriate suggestions. Future work should involve prospective clinical validation to assess the real-world impact of K-NeSyNet on treatment outcomes.

## 6. Conclusion

In this paper, we presented K-NeSyNet, a Knowledge-driven Neuro-Symbolic Network for personalized oncology treatment recommendation. By integrating a comprehensive Multi-Modal Oncology Knowledge Graph (MM-OKG) with a three-channel differentiable symbolic reasoning module and an adaptive gated fusion network, K-NeSyNet effectively bridges the gap between data-driven deep learning and rule-based clinical decision support. Extensive experiments on a real-world dataset of 4,781 oncology patients demonstrated that K-NeSyNet consistently outperforms eight state-of-the-art baselines on top-K recommendation metrics while maintaining competitive clinical safety performance. The ablation study confirmed the critical role of the adaptive fusion gate, whose removal caused the most significant performance degradation. Furthermore, the model’s ability to generate transparent, score-decomposed explanations provides a crucial step toward building physician trust in AI-driven clinical tools.

Future work will focus on three directions: (1) expanding the MM-OKG to include longitudinal patient data, such as temporal changes in tumor characteristics and sequential treatment responses; (2) enriching the contraindication knowledge base with more granular, dose-dependent adverse interaction data; and (3) conducting prospective clinical validation studies to assess the real-world utility of K-NeSyNet as a clinical decision support tool.

## Data Availability

All data used in this study are publicly available from open databases. TCGA data can be accessed via NCI Genomic Data Commons (https://gdc.cancer.gov/). DGIdb (https://www.dgidb.org/) and KEGG (https://www.genome.jp/kegg/) are publicly accessible online databases.

https://www.dgidb.org

https://www.genome.jp/kegg

## CRediT authorship contribution statement

**Liuqing Yang:** Investigation, Methodology, Writing – original draft. **Hong Wan:** Data curation, Formal analysis. **Jiangping Zhu:** Software, Validation. **Pei Zhou:** Visualization. **Zhongjian Wang:** Conceptualization, Supervision, Writing – review & editing.

## Data availability

All data used in this study are from publicly available, de-identified datasets. TCGA data are available through the NCI Genomic Data Commons (GDC). DGIdb and KEGG data are publicly accessible research databases. The code and constructed knowledge graph will be made available upon acceptance.

## Acknowledgments

This research was supported by the Internal Artificial Intelligence Fund of BCPM Data Limited. The authors thank the anonymous reviewers for their constructive feedback.

